# Acute cognitive deficits after traumatic brain injury predict Alzheimer’s disease-like degradation of the human default mode network

**DOI:** 10.1101/2020.08.01.20166561

**Authors:** Andrei Irimia, Alexander S. Maher, Nikhil N. Chaudhari, Nahian F. Chowdhury, Elliot B. Jacobs, the Alzheimer’s Disease Neuroimaging Initiative

## Abstract

Traumatic brain injury (TBI) and Alzheimer’s disease (AD) are prominent neurological conditions whose neural and cognitive commonalities are poorly understood. The extent of TBI-related neurophysiological abnormalities has been hypothesized to reflect AD-like neurodegeneration because TBI can increase vulnerability to AD. However, it remains challenging to prognosticate AD risk partly because the functional relationship between acute post-traumatic sequelae and chronic AD-like degradation remains elusive. Here, functional magnetic resonance imaging (fMRI), network theory and machine learning (ML) are leveraged to study the extent to which geriatric mild TBI (mTBI) can lead to AD-like alteration of resting-state activity in the default mode network (DMN). This network is found to contain modules whose extent of AD-like, post-traumatic degradation can be accurately prognosticated based on the acute cognitive deficits of geriatric mTBI patients with cerebral microbleeds. Aside from establishing a predictive physiological association between geriatric mTBI, cognitive impairment and AD-like functional degradation, these findings advance the goal of acutely forecasting mTBI patients’ chronic deviations from normality along AD-like functional trajectories. The association of geriatric mTBI with AD-like changes in functional brain connectivity as early as ∼6 months post-injury carries substantial implications for public health because TBI has relatively high prevalence in the elderly.

## Introduction

Traumatic brain injury (TBI) can result in functional brain alterations causing neural and cognitive deficits [1, 2]. Even after mild TBI (mTBI), any cognitive domain can be affected by such deficits [3], whose manifestation may accelerate the onset of mild cognitive impairment (MCI) [4, 5], particularly in geriatric patients [6]. Although neurotrauma increases the risk for Alzheimer’s disease (AD) [7], there is little knowledge on how TBI affects functional trajectories and gives rise to neuropathophysiology. Furthermore, the relationship between TBI severity and AD-like brain dysfunction is poorly understood, and the mechanisms whereby TBI can elicit functional abnormalities which increase AD risk remains unclear. One hypothesis is that post-traumatic functional changes exhibit patterns which progressively resemble those of AD [8, 5, 9]; if this is the case, characterizing AD-like brain alterations after TBI could assist in improving AD risk assessment and the early identification of TBI patients at higher risk for this disease.

The default mode network (DMN) is a large-scale brain network which is most commonly active when a person is at wakeful rest [10]. In this resting state (RS), individuals lie quietly awake without performing tasks or being exposed to stimuli, and their DMN activity can be recorded using techniques like functional magnetic resonance imaging (fMRI). RS fMRI recordings only require passive participation from study participants, such that the prospect of isolating AD prognosticators from such data is logistically and clinically appealing. Although DMN alterations have been quantified in both AD and TBI [11-13], whether and how post-traumatic DMN abnormalities reflect AD-like functional degradation remains unknown. Because the DMN includes some of the longest white matter tracts in the brain (including tracts which integrate brain activity across the corpus callosum) [14], this network is particularly vulnerable to diffuse axonal injury after trauma [15]. Thus, studying TBI-related alterations in the DMN of subjects with cerebral microbleeds (CMBs, which are biomarkers of non-focal axonal injury [16])—as opposed to changes in other, less broadly-distributed networks—is attractive. To identify post-traumatic biomarkers of AD risk, focusing on mTBI is appealing because the mTBI population is considerably larger, more homogeneous, and logistically easier to study than that of patients with moderate-to-severe TBI. Additionally, geriatric TBI is a promising setting for studying how this condition can lead to AD-like neural degradation because the comparison of AD patients to young or middle-aged TBI patients is confounded by aging effects.

This study leverages fMRI recordings acquired from healthy controls (HCs), geriatric mTBI participants with CMBs and AD patients to provide evidence that, within ∼6 months post-mTBI, functional connectivity (FC) within the DMN exhibits deviations from normality whose spatiotemporal properties are statistically indistinguishable from those of similar deviations observed in AD. A linear combination of *acute* post-traumatic cognitive scores is found to be significantly and sensitively associated with *chronic* AD-like RS DMN alterations. Additionally, a supervised machine learning (ML) classifier is found to accurately identify mTBI patients with relatively broad *chronic* abnormalities in the DMN based on *acute* cognitive performance. These findings establish a detailed functional and connectomic relationship between mTBI-related acute cognition and AD-like DMN features, whose further characterization may facilitate the early identification of geriatric mTBI patients with CMBs at relatively high risk for AD.

## Methods

### Participants

This study was conducted with Institutional Review Board approval. Included were one HC group (*N*_1_ = 48, 22 females; age: µ = 69 y, σ = 5 y, range: 58-79 y), and two study groups: geriatric mTBI (*N*_2_ = 29, 13 females; age: µ = 68 y, σ = 6 y; range: 57-79 y) and AD (*N*_3_ = 37, 19 females; age: µ = 70 y, σ = 8 y; range: 55-84 y). HC and AD subjects were selected from the AD Neuroimaging Initiative (ADNI) cohort, whose eligibility criteria are described elsewhere [17]. A total of 15 HC volunteers (31%), 9 TBI participants (38%) and 17 AD patients (46%) were hypertensive. Some ADNI participants were receiving hormonal treatment (HC: *N* = 14 or 29%; AD: *N* = 9 or 24%); some were taking medications for neurological and/or psychiatric disease (HC: *N* = 20 or 42%; TBI: *N* = 18 or 62%; AD: *N* = 36 or 97%), vascular disease (HC: *N* = 29 or 60%; TBI: *N* = 19 or 65%; AD: *N* = 22 or 59%), or metabolic disease (HC: *N* = 6 or 13%; TBI: *N* = 3 or 10%; AD: *N* = 5 or 14%). To be included, all participants had to have Montréal Cognitive Assessment (MoCA) scores and a complete session of RS fMRI data. HC participants had been clinically evaluated as having normal cognition; their MoCA scores ranged from 22 to 29 (µ = 26, σ = 2). AD patients’ scores ranged from 6 to 25 (µ = 17, σ = 5), and all had a clinical AD diagnosis; TBI participants’ scores were acquired within 48 hours post-injury and were between 20 and 29 (µ = 23, σ = 2). Mini Mental State Examination (MMSE) scores were available for both HC (µ = 29, σ = 1; range: 26 to 30), TBI volunteers (µ = 22, σ = 7; range: 13 to 29) and AD participants (µ = 20, σ = 5; range: 9 to 28). AD patients had Clinical Dementia Rating (CDR) sub-scores between 2 and 17 (µ = 6, σ = 4), whilst HCs had CDR sub-scores between 0 and 2 (µ = 1, σ = 0.7); CDR scores were not available for TBI participants. For HCs, the number of apolipoprotein E (ApoE) ε4 alleles was zero for 54% of the sample, one for 31% and two for 15%. For AD patients, 24% had no ε4 alleles, 46% had one and 30% had two. No ApoE allele information was available for TBI participants. TBI volunteers had fMRI recordings acquired ∼6 months post-injury (µ = 5.6 months, σ = 0.5 months) at 3 T, i.e. the same scanner field strength as the HC and AD participants. They had to have (a) a TBI due to a fall, (b) no clinical findings on acute *T*_*1*_/*T*_*2*_-weighted MRI, (c) no clinical findings other than CMBs on susceptibility weighted imaging (SWI), (d) an acute Glasgow Coma Scale score greater than 12 (µ = 13.7, σ = 0.5) upon initial medical examination, (e) loss of consciousness of fewer than 30 minutes (µ ≃ 4 minutes, σ ≃ 8 minutes), (f) post-traumatic amnesia of fewer than 24 hours (µ ≃ 3.5 hours, σ ≃ 3.2 hours), and (g) a lack of clinical history involving pre-traumatic neurological disease, psychiatric disorder or drug/alcohol abuse. CMBs were identified in each subject using an automatic algorithm for CMB segmentation [18] and the validity of the findings were confirmed by two human experts with training in CMB identification from SWI, who had been blinded to automatic segmentation results. Disagreements between these experts were resolved by a third one (AI). Null hypotheses of group differences in age and cognition were tested using Welch’s two-tailed *t* test for samples with unequal variances. The null hypothesis of independence between sex and group membership was tested using Pearson’s *χ*^2^ test. Effect sizes were quantified using Cohen’s *d* for Welch’s *t* test and the *ϕ* coefficient for Pearson’s *χ*^2^ test.

### Neuroimaging

HC and AD participant data used in the preparation of this article were obtained from the Alzheimer’s Disease Neuroimaging Initiative (ADNI) database (http://adni.loni.usc.edu). ADNI was launched in 2003 as a public-private partnership, led by Principal Investigator Michael W. Weiner, MD. The primary goal of ADNI has been to test whether serial MRI, positron emission tomography (PET), other biological markers, and clinical and neuropsychological assessment can be combined to measure the progression of MCI and early AD. For up-to-date information, see www.adni-info.org. fMRI volumes were acquired at 3 T using the ADNI acquisition protocol [19]. An average of 140 fMRI volumes were obtained using the following parameters: *T*_*R*_ = 3 s; *T*_*E*_ = 30 ms; flip angle = 80°; slice thickness ≃ 3.3 mm; 48 slices). TBI subjects’ fMRI data were acquired in a Siemens Trio TIM 3 T scanner using an acquisition protocol very similar to the ADNI protocol.

### Preprocessing

fMRI analysis was implemented using the Free Surfer (FS) Functional Analysis Stream (FS-FAST, https://surfer.nmr.mgh.harvard.edu/fswiki/FsFast) with default parameters for (a) motion correction, (b) frame censoring, (c) frequency filtering, (d) brain masking, (e) intensity normalization, (f) co-registration of fMRI volumes to *T*_*1*_-weighted volumes, (g) surface sampling to the FS atlas, (h) smoothing (kernel with full width of 5 mm at half maximum), (i) surface and volume sampling to the Montreal Neurological Institute (MNI) atlas containing 305 subjects, and (j) smoothing for subcortical structure analysis. The first four volumes in each fMRI time series were discarded to preserve signal equilibrium and to account for each participant’s adaptation to the sequence; the rest were used for analysis [20]. Nuisance variables (cerebrospinal fluid, white matter, and motion correction parameters) were accounted for using FS-FAST.

### fMRI seeds

Seeds were derived in a two-step process. In the first step, the cortical delineation of the DMN defined by Yeo et al. [14] was used to parcel the cortex. This delineation includes the following cortical regions: (a) *frontal* (prefrontal cortex, precentral ventral cortex, anterior cingulate cortex, etc.), (b) *medial temporal/retrosplenial* (the parahippocampal complex), (c) *lateral temporal* (the inferior temporal gyrus and superior temporal sulcus), (d) *lateral parietal* (inferior parietal, intraparietal regions, etc.) and (e) *medial parietal/posterior cingulate* (posterior cingulate cortex and part of the precuneus). In the second step, Yeo regions were divided into gyral/sulcal parcels based on the intersection of the Yeo regions with the cortical parcellation scheme of Destrieux et al. [21]. In other words, the final set of fMRI seeds consisted of all the regions which resulted from the intersection of the Yeo and Destrieux schemes. This was deemed to provide greater spatial detail to the analysis, particularly for DMN regions with substantial cortical coverage (e.g. the frontal DMN, which includes a large contiguous portion of cerebral cortex). The intersection of the Destrieux and Yeo schemes led to the delineation of 46 DMN regions (22 cortical regions and the hippocampus for each hemisphere; see caption to **Figure 1**).

**Figure 1.**
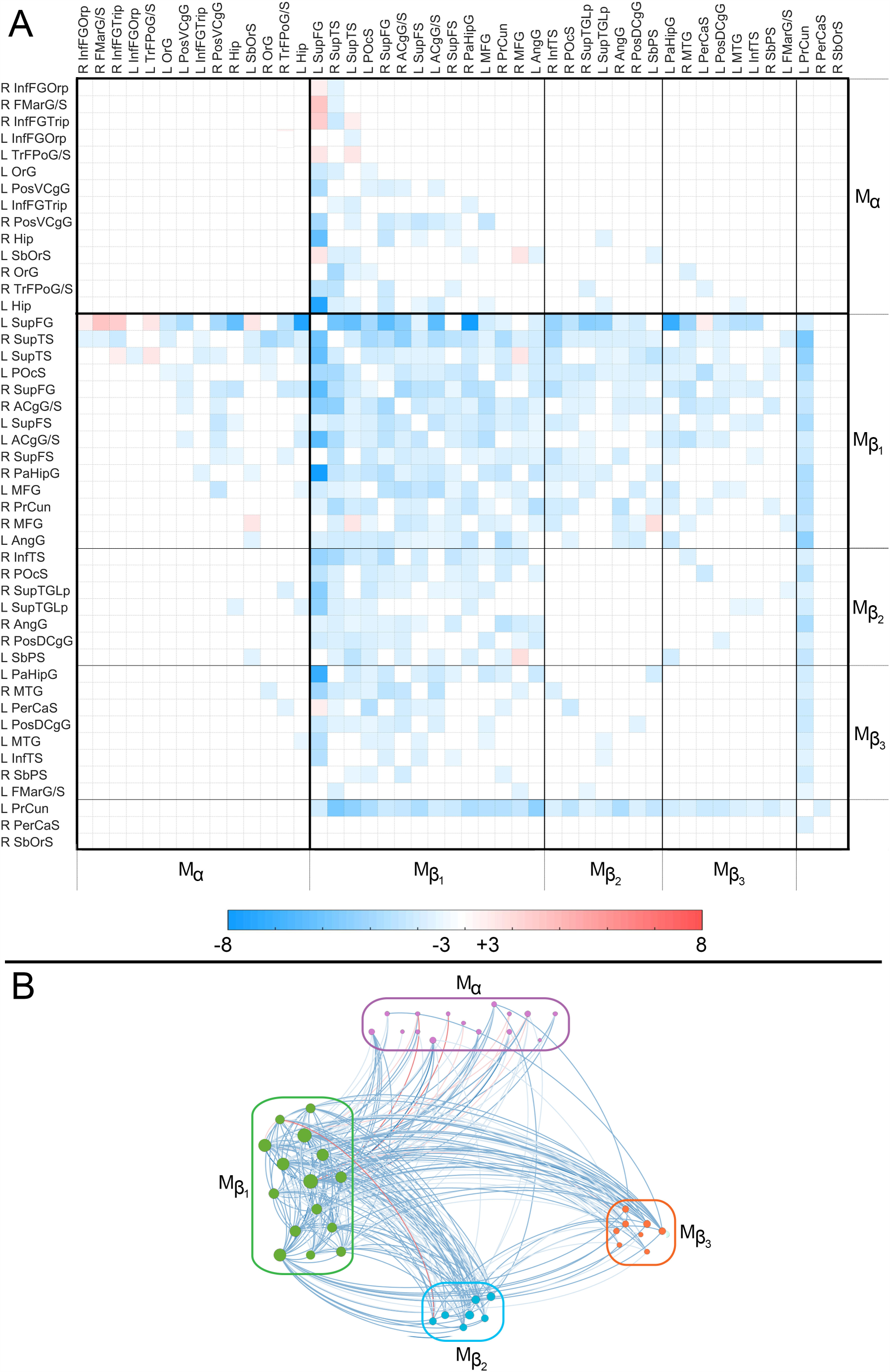
(A) The dissimilarity matrix *D*(*HC, TBI*) displays significant differences in the FC *ρ* between the HC and mTBI groups. Each cell *D*_*ij*_(*HC, TBI*) encodes the result of testing the null hypothesis of no mean difference in *ρ*_*ij*_ between groups. Cells corresponding to region pairs for which the null hypothesis fails to be rejected are drawn in white. Elsewhere, the color-coded quantity is a *t* statistic with 75 *df*. If *ρ*_*ij*_(*TBI*) > *ρ*_*ij*_(*HC*), *t* is positive and *D*_*ij*_ is drawn in red; otherwise, *t* is negative and *D*_*ij*_ is drawn in blue. Boundaries between *M*_*α*_ and *M*_*β*_ are delineated by thick black lines; boundaries between the submodules of *M*_*β*_ are delineated by thinner lines. Regions are labeled using the connectogram abbreviations of Irimia et al. [65]. Frontal regions are the FMarG/S, InfFGOrp, InfFGTrip, MFG, OrG, SbOrS, SupFG, SupFS, and TrFPoG/S; limbic regions are the ACgG/S, PerCaS, PosDCgG, and PosVCgG; temporal regions are the InfTS, PaHipG, SupTGLp, and SupTS; parietal regions are the AngG, POcS, PrCun, and SbPS. (B) Graph representation of *D*(*HC, TBI*). Nodes are color-coded and grouped by module. Edge colors encode *t* score values, according to the color bar in (A). Abbreviations: R = right; L = left; Hip = hippocampus. Cortical region abbreviations: ACgG/S = anterior cingulate gyrus and sulcus; AngG = angular gyrus; FMarG/S = frontomarginal gyrus and sulcus; InfFGOrp = inferior frontal gyrus, orbital part; InfFGTrip = inferior frontal gyrus, triangular part; InfTS = inferior temporal sulcus; MFG = middle frontal gyrus; OrG = orbital gyrus; PaHipG = parahippocampal gyrus; PerCaS = pericallosal sulcus; POcS = parieto-occipital sulcus; PosDCgG = posterior dorsal cingulate gyrus; PosVCgG = posterior ventral cingulate gyrus; PrCun = precuneus; SbOrS = suborbital sulcus; SbPS = subparietal sulcus; SupFG = superior frontal gyrus; SupFS = superior frontal sulcus; SupTGLp = superior temporal gyrus, lateral part; SupTS = superior temporal sulcus; TrFPoG/S = transverse frontopolar gyrus and sulcus.

### FC analysis

Both within each subject’s space and within MNI space, individual fMRI time series *s* were consolidated using `isxconcat-sess`. A weighted least-squares (WLS) general linear model (GLM) was implemented using mri_glmfit to identify pairs of anatomic regions (*i, j*) whose fMRI signals *s*_i_ and *s*_j_ had a statistically significant partial correlation *ρ*_*ij*_. For each DMN seed parcel *i*, a subject-level, voxel-wise analysis was implemented to identify spatially contiguous clusters in target region *j* (*i* ≠ *j*) within which *ρ*_*ij*_ is significant. How *ρ*_*ij*_ differed in each study group (TBI or AD) relative to HC was investigated using the same GLM. Effect sizes were quantified using Cohen’s *f*^2^.

### Equivalence testing

One key premise of this study is that brain features which are both (a) significantly different from HCs in both TBI and AD, *and* (b) significantly similar across TBI and AD, can be said to be *AD-like*. Thus, a brain feature observed in TBI patients can be said to be AD-like if the feature in question differs from HCs in both TBI and AD, *and* is also significantly similar across *both* TBI and AD. If *ρ*_*ij*_ differs significantly from HCs in both TBI *and* AD, a null hypothesis of *statistical equivalence* can be tested to determine whether *ρ*_*ij*_ is AD-like. Formally, for two samples *A* and *B*, a null hypothesis of equivalence can be stated as µ_*A*_(*ρ*_*ij*_) ≠ µ_*B*_(*ρ*_*ij*_) rather than as the conventional null hypothesis µ_*A*_(*ρ*_*ij*_) = µ_*B*_(*ρ*_*ij*_). The null hypothesis of equivalence fails to be accepted if the two means fall within the interval (−*δ, δ*), where *δ* is the equivalence margin of the test [22]. In statistical parlance, equivalence implies that the correlations *ρ*_*ij*_(*A*) and *ρ*_*ij*_(*B*) are sufficiently close that neither can be considered greater or smaller than the other [23]. Equivalence hypotheses can be tested using two one-sided *t* tests (TOSTs) [24]; in this study, *δ* is assigned a conservative value equal to 0.2 times the width of the 95% confidence interval for the difference µ_*A*_(*ρ*_*ij*_) − µ_*B*_(*ρ*_*ij*_). To identify TBI and AD participants’ correlations which deviate appreciably from normality (i.e. from the HC group) in both former groups, the null hypothesis of equivalence is only tested if both µ_*TBI*_(*ρ*_*ij*_) and µ_*AD*_(*ρ*_*ij*_) differ significantly from µ_HC_(*ρ*_*ij*_). Effect sizes were quantified using Cohen’s *f*^2^. Multiple comparison corrections using 300 permutations and a cluster-wise *p*-value threshold of 0.05 were implemented for all statistical tests. Equivalence testing was implemented using freely available MATLAB software (https://www.mathworks.com/matlabcentral/fileexchange/63204).

### (Dis)similarity matrices

Two *dissimilarity matrices D*(*TBI, HC*) and *D*(*AD, HC*) were assembled to describe mean differences in *ρ*_*ij*_ between HCs and each of the study groups (TBI and AD), respectively. Each matrix element *D*_*ij*_(*TBI, HC*) is set to the value of the *t* statistic for the test of the null hypothesis *ρ*_*ij*_(*TBI*) − *ρ*_*ij*_(*HC*) = 0, and a similar procedure is used for *D*_*ij*_(*AD, HC*). A *similarity matrix S*(*TBI, AD*) was also calculated to describe significant statistical equivalences of *ρ*_*ij*_ across study groups. Each matrix element *S*_*ij*_ is set to the TOST *t* statistic which had the smallest magnitude.

### Network analysis

To investigate DMN-related commonalities and differences between TBI and AD, three analyses were carried out. The first two involved studying *D*(*HC, TBI*) and *D*(*HC, AD*) to group DMN nodes based on how TBI modulated their correlation differences relative to HCs and ADs, respectively. The third one examined *S*(*TBI, AD*) to identify DMN nodes affected equivalently in both TBI and AD. To identify network modules, the Louvain algorithm for community detection [25] was applied 100 times for each matrix to identify module partitions. The symmetric reverse Cuthill-McKee (RCM) ordering [26] of each module was then calculated to rearrange nodes within each module. This method permutes the rows and columns of a symmetric sparse matrix to form a band matrix with minimal bandwidth, i.e. whose nonzero elements are optimally close to the diagonal. The algorithm identifies a pseudo-peripheral vertex of the network, and then utilizes a breadth-first search to order vertices by decreasing distance from the pseudo-peripheral vertex. When applied to each module of *S*, such blocks are arranged along the main diagonal and produce a visual representation which facilitates module inspection and analysis. Network analysis was implemented using the freely available Brain Connectivity Toolbox (sites.google.com/site/bctnet).

### Network randomization

To determine whether modules’ node memberships were dependent upon the DMN parcellation scheme used in the study, the DMN was reparcelled randomly 100 times to generate alternative parcellations which had the same number of nodes as the original DMN but different cortical patches corresponding to each node. An approach similar to those of Gordon et al. [27] and Irimia&Van Horn [28] was used to obtain randomized parcellations of the DMN. Briefly, random points within the cortical coverage of the DMN were selected. From these seeds, parcels were simultaneously expanded outward on the cortical mesh until they met either other parcels or the boundary of the DMN. The procedure for identifying network modules was implemented for each randomized parcellation and the modularity structure of the network was found each time by applying the Louvain algorithm 100 times to each matrix. The spatial overlap between each original module and the randomized modules was quantified using the Sørensen-Dice coefficient [29].

### Acute cognitive impairment vs. chronic brain function

A multivariate regression analysis was implemented to study the relationship between TBI patients’ *acute* MoCA scores and the number of their *chronic* FCs which were statistically equivalent to those of AD patients. The latter involved region pairs with the largest absolute values of *S*_*ij*_ (highest similarity across TBI and AD): (a) the right superior temporal sulcus and the right anterior cingulate gyrus/sulcus, (b) the left and right superior frontal gyri, (c) the left hippocampus and superior temporal sulcus, and (d) the left middle frontal gyrus and the ventral part of the posterior cingulate gyrus. The predictor variables were the entries in *S* associated with these region pairs, and the response variable was the MoCA score. Sex, age at MRI acquisition and educational attainment were included as covariates. Cohen’s *f*^2^ was used as a measure of effect size and the null hypothesis of overall regression was tested using Fisher’s *F* test [30]. To confirm and to broaden regression findings, a support vector machine (SVM) was implemented in MATLAB (http://mathworks.com) with default parameters and using the iterative single data algorithm (ISDA), a linear kernel function and a heuristically assigned kernel scale parameter. The SVM was trained and cross-validated ten-fold to distinguish (a) TBIs with relatively *moderate* AD-like DMN deviations from normality (i.e. with 7 or fewer statistical equivalences across TBI and AD) from other TBIs and also (b) TBIs with relatively *extensive* abnormalities (i.e. with at least 15 equivalences) from other TBIs. Let *N*_*E*_ be the number of significant equivalences identified (*N*_*E*_ = 22 here, see **Figure 3**). Then the thresholds values of 7 and 15 correspond to ⌊*N*_*E*_/3⌋ and ⌈2*N*_*E*_/3⌉, respectively (**Figure 3**). For the SVM, the number of true negatives (TNs), true positives (TPs), false negatives (FNs) and false positives (FPs) were computed, as were the true positive rate (TPR, or sensitivity), true negative rate (TNR, or specificity), positive prediction value (PPV, or precision) and Matthews’ correlation coefficient (MCC) [31]. Regression and SVM analyses were implemented in MATLAB using the glmfit, fitcsvm and predict functions.

### Visualization

Matrices were visualized to identify and examine DMN modules (**Figure 1, Figure 2**, and **Figure 3**). Matrix entries were thresholded by statistical significance; for example, if *ρ*_*ij*_ does not differ significantly between the groups compared, the cell for *D*_*ij*_ is drawn in white. Similarly, if *ρ*_*ij*_ does not differ significantly between TBI and AD, the cell for *S*_*ij*_ is also drawn in white. For statistically significant results, dissimilarity matrix cells are drawn in either red or blue, depending on their sign (see the caption to **Figure 3**). To facilitate inspection, the cortical regions within each module were drawn on an average atlas representation of the brain. Graph representations of each functional correlation matrix were also generated using Gephi software (http://gephi.org). In these, each region’s node was depicted as a circle whose diameter was proportional to the number of cortical regions to which the region represented by the node was functionally connected. Similarly, edges were colored using shades of blue or red to reflect the *t* score of lowest magnitude associated with the TOSTs for statistical equivalence testing.

**Figure 2.**
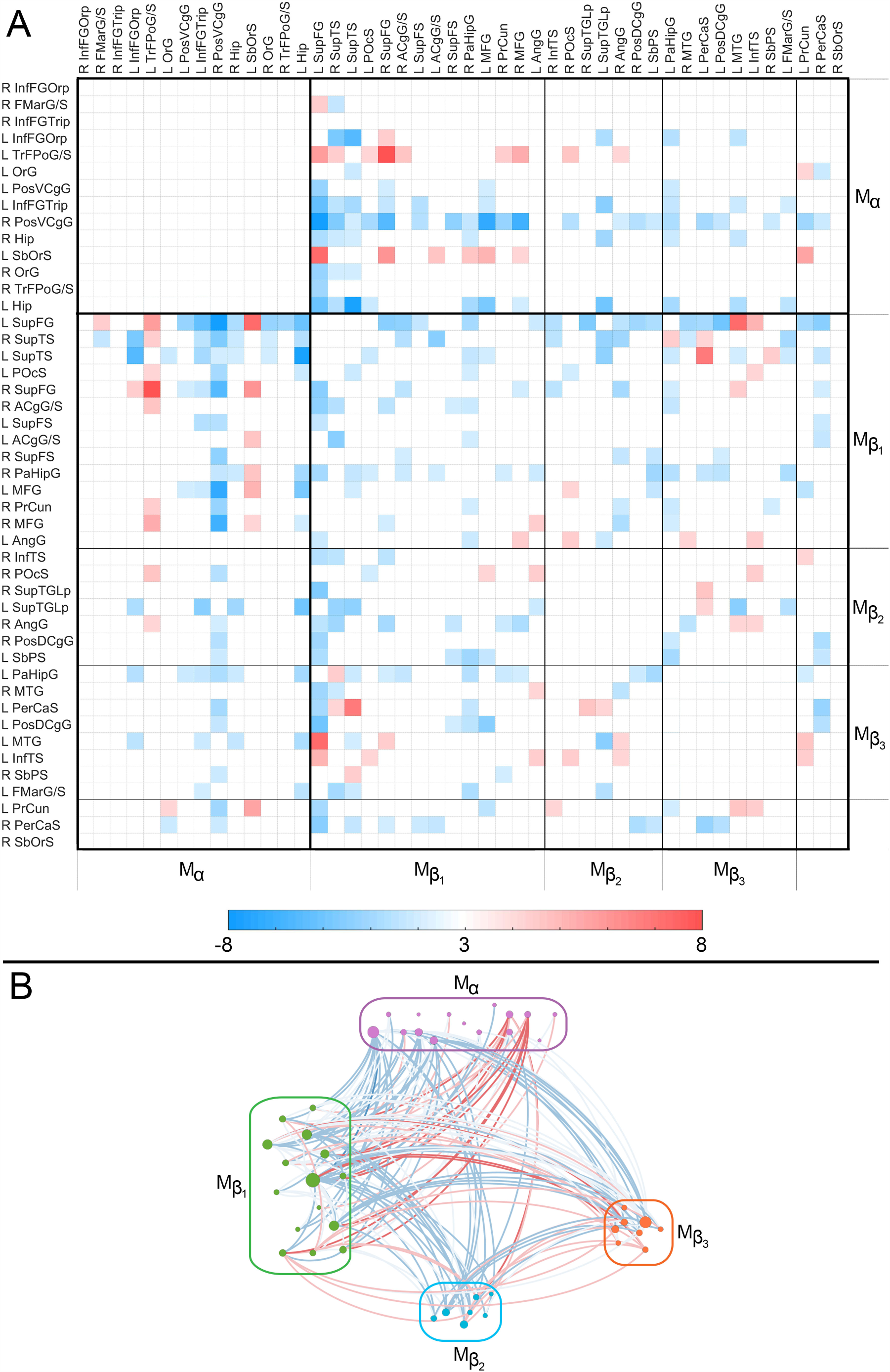
Like Figure 1, for *D*(*HC, AD*). The color-coded quantity is a *t* statistic with 82 *df*. See the caption of Figure 1 for abbreviations.

**Figure 3.**
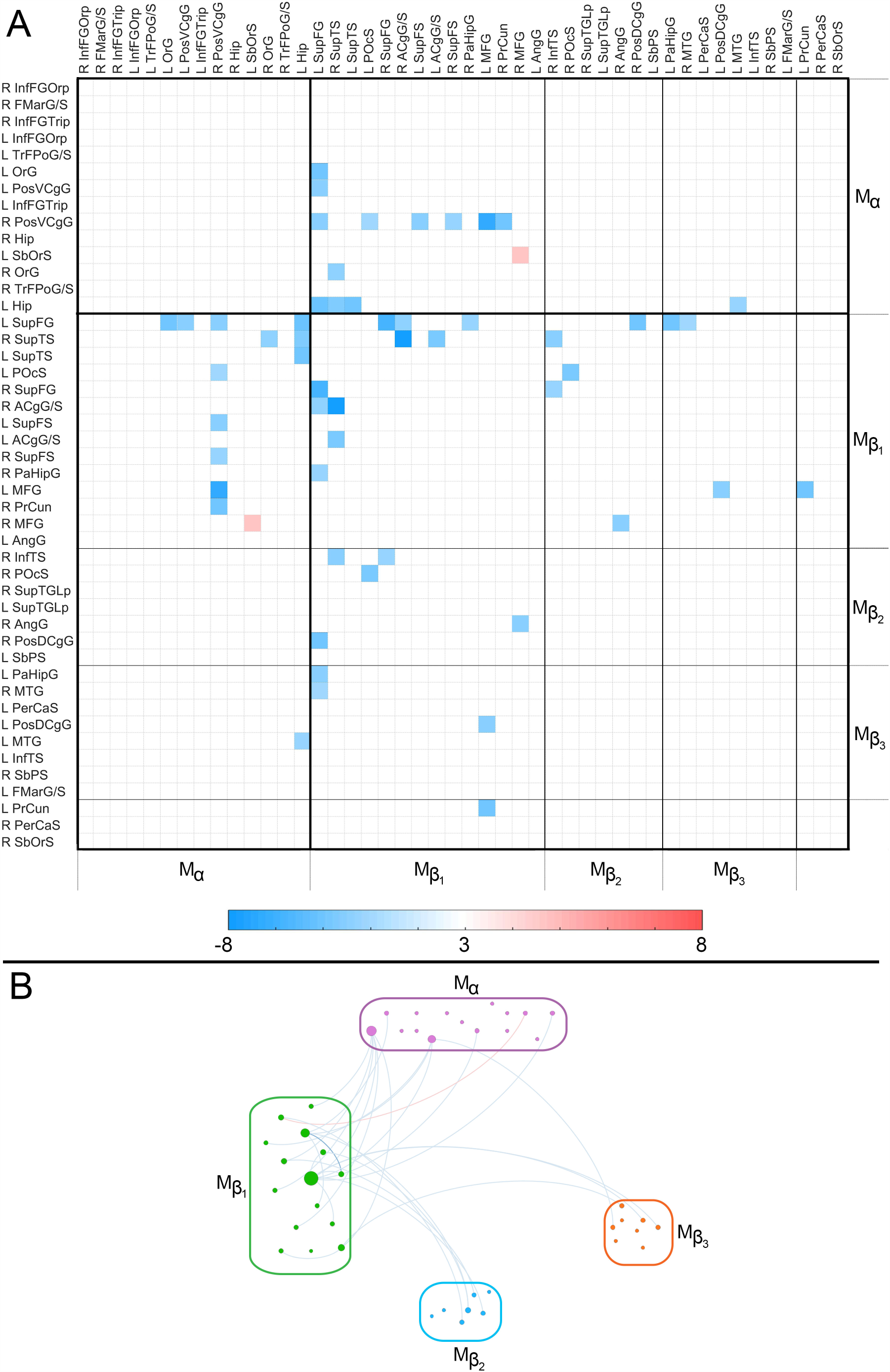
The similarity matrix *S*(*TBI, AD*) displays statistically significant equivalences of functional correlation *ρ* between the TBI and AD groups. Each cell *S*_*ij*_ encodes the result of testing the null hypothesis of equivalence in *ρ*_*ij*_ across these two groups. This null hypothesis is only tested if both TBI and AD differ significantly from HC. In other words, if there is no significant mean difference in *ρ* between either (a) HC and TBI *and/or* (b) HC and AD, *S*_*ij*_ is drawn in white. If both study groups differ from the HC group but no significant statistical equivalence is found across TBI and AD, *S*_*ij*_ is also drawn in white. Elsewhere, the color-coded quantity is the value of the TOST procedure’s *t* statistic with the smallest magnitude. The color-coded quantity is a *t* statistic with 63 *df*. If a statistical equivalence is associated with a relatively stronger correlation in both TBI and AD relative to HC, *t* is positive and *S*_*ij*_ is drawn in red; if the correlation is weaker relative to HC, *t* is negative and *S*_*ij*_ is drawn in blue. Boundaries between *M*_*α*_ and *M*_*β*_ are delineated by thick black lines; boundaries between the submodules of *M*_*β*_ are delineated by thinner lines. See the caption of Figure 1 for abbreviations. (B) Graph representation of *S*(*TBI, AD*). Nodes are color-coded and grouped by module. Edge colors encode *t* score values, according to the color bar in (A).

## Results

### Demographics

Three cohorts were studied: HC participants (48 subjects, 22 females; age µ ± σ = 69 ± 5 years (y)), geriatric mTBI subjects with CMBs (29 subjects, 13 females; 68 ± 6 y) and AD patients (37 subjects, 18 females; 74 ± 8 y). Further demographic descriptors are provided in the Methods section. CMB counts were found to range from 0 to 43 (µ ± σ = 13 ± 9) in HCs, from 0 to 89 (µ ± σ = 17 ± 14) in mTBI volunteers, and from 0 to 6 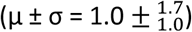 in AD patients. No significant differences in mean age were found between HC and TBI participants (*t* = 0.52, *df* = 47, *p* = 0.60, Cohen’s *d* = 0.36), between HC and AD volunteers (*t* = −0.95, *df* = 55, *p* = 0.17, Cohen’s *d* = 0.16) or between TBI and AD patients (*t* = −1.24, *df* = 64, *p* = 0.11, Cohen’s *d* = 0.46). No significant differences in sex ratios were found across groups (*χ*^2^ = 0.22, *df* = 1, *p* = 0.90, *ϕ* = 0.09). Significant differences in MoCA scores were found between HC and acute TBI participants (*t* = 5.7, *df* = 50, *p* < 0.001, Cohen’s *d* = 1.50), between HC and AD participants (*t* = 9.0, *df* = 44, *p* < 0.001, Cohen’s *d* = 2.36) but not between acute TBI participants and AD patients (*t* = 0.4, *df* = 61, *p* = 0.65, Cohen’s *d* = 1.58). Similarly, significant differences in MMSE scores were found between HC and acute TBI participants (Welch’s *t* = 5.4, *df* ≃ 28.7, *p* < 0.001, Cohen’s *d* = 1.2), between HC and AD participants (Welch’s *t* = 10.8, *df* ≃ 37.2, *p* < 0.001, Cohen’s *d* = 2.7) but not between acute TBI participants and AD patients (Welch’s *t* = 1.3, *df* ≃ 48.7, *p* = 0.1, Cohen’s *d* = 0.3).

### (Dis)similarity matrices and modularity

Participants’ DMNs were delineated and then parceled into gyri and sulci based on the morphometric boundaries between cortical structures, as previously described. For each pair of regions *i* and *j*, clusters of significant functional correlations *ρ*_*ij*_ were then identified. Two *dissimilarity matrices D*(*TBI, HC*) and *D*(*AD, HC*) were computed to quantify significant mean differences in *ρ*_*ij*_ between HC and each of the study groups (TBI and AD, respectively). A *similarity matrix S*(*TBI, AD*) was also calculated to describe significant statistical equivalences of *ρ*_*ij*_ across TBI and AD. Both similarity and dissimilarity were quantified using Student’s *t* scores (see Methods).

To determine which brain regions are similarly vulnerable to TBI-and to AD-related deviations from normality (i.e. from HCs), network modules were identified within each (dis)similarity matrix. For reproducibility, the dependence of module composition upon the anatomy-based parcellation scheme was also explored. This was done by repartitioning the DMN randomly and repeatedly to create alternative parcellations which had the same number of nodes—but different spatial configurations—as the original, anatomy-based parcellation. The process of identifying network modules was then repeated for each of these randomized parcellations. At every iteration, DMN modules were identified in each dissimilarity matrix; the number *N*_*R*_ of randomized modules (µ ± σ = 2.01 ± 0.3) was not found to differ significantly from the number of modules *N*_*R*_ obtained using the anatomic parcellation (*N*_*A*_= 2; Student’s *t* > 0.37, *df* = 99, Cohen’s *d* = 0.03). Furthermore, the original and randomized modules overlapped spatially with high consistency across the 100 randomizations (Sørensen-Dice coefficient µ ± σ = 0.94 ± 0.06; 95% CI = [92.89, 95.10]). Thus, the randomized modules’ nodal memberships and spatial coverages agreed with those of the original modules.

### DMN modules in mTBI versus HC

RCM orderings [26] were used to display modules along the main diagonal of each (dis)similarity matrix. The RCM-ordered modules of *D*(*HC, TBI*) and *D*(*HC, AD*) are displayed in **Figure 1** and **Figure 2**, respectively, as are the graph representations of the corresponding dissimilarity matrices. *M*_*α*_, the first module common to both *D*(*HC, TBI*) and *D*(*HC, AD*), includes (a) the hippocampus, (b) ventral and dorsal prefrontal cortex and (c) the ventral aspect of the posterior cingulate gyrus. Significant functional correlations between nodes within *M*_*α*_ do not differ significantly across TBI and HC (**Figure 1**); this may indicate that, on average, the geriatric mTBI patients did not experience substantial FC alterations within brain areas covered by *M*_*α*_ within the first ∼6 months post-injury. *M*_*β*_, the second module along the diagonal of *D*(*HC, TBI*), contains all the DMN regions outside *M*_*α*_. Within *M*_*β*_, correlations are consistently weaker in mTBI participants relative to HCs. Furthermore, the superior frontal gyrus and precuneus are found to be the two structures whose TBI-related FC deviations from normality are broadest across the DMN. FC differences between TBI and HC involve relatively few pathways connecting *M*_*α*_ and *M*_*β*_. Thus, although *M*_*α*_ connections are considerably less affected by TBI compared to *M*_*β*_, some pathways between these modules are not.

### DMN modules in AD versus HC

Whereas FC is typically weaker in TBI than in HC (**Figure 1**), AD patients’ FC deviations from normality vary considerably (**Figure 2**). These deviations can be grouped into five modules which occur bilaterally; the first is identical to *M*_*α*_ and the rest are subdivisions of *M*_*β*_ (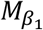 through 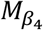). Across AD and TBI, 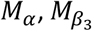 and 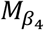 are similar in that their intramodular FCs do not differ significantly from those of HC participants. 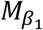 is a fronto-temporal module with relatively few intramodular FC differences between AD and HC, but with considerably more differences of this kind involving intermodular connections. 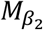 contains broadly distributed frontal, limbic and parietal regions; comparing the HC and AD groups from the standpoint of correlations within 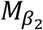 reveals sparse group differences involving both intra-and inter-modular connections. Like in the case of *M*_*α*_, AD patients’ 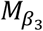 and 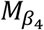 modules do not have intramodular correlations which differ significantly from HC, although both subunits exhibit numerous such differences involving intermodular connections.

### DMN modules in TBI versus AD

**Figure 3** displays the similarity matrix *S*(*TBI, AD*) and its graph representation to identify regions with statistical equivalences across study groups. Regions exhibiting such similarities involve dorsolateral prefrontal cortex, the lateral temporal lobe, and the ventral aspect of the posterior cingulate gyrus. The hippocampus is topologically proximal to the latter two areas; furthermore, hippocampo-cortical correlations are substantially affected in both TBI and AD. Nevertheless, only few hippocampo-cortical correlations are statistically equivalent across these conditions. The strongest similarities between TBI and AD involve connectivity between the lateral temporal lobe and anterior cingulate cortex, as well as between dorsolateral prefrontal cortex and each of the following three structures: the hippocampus, the lateral temporal lobe, and posterior cingulate cortex.

### Acute cognition versus chronic DMN in TBI

Upon testing the association between MoCA scores and the number of FC similarities (i.e. pairwise statistical equivalences) between TBI and AD, the null hypothesis of the test for overall multivariate regression was rejected (*F* = 1.6, *df*_1_ = 5, *df*_2_ = 111, *p* < 0.0034, Cohen’s *f*^2^= 0.39). In other words, this test rejected the null hypothesis according to which there was no multivariate correlation between (a) MoCA scores and (b) the number of FC similarities involving TBI and AD. To study further the relationship between acute cognition and chronic DMN dysfunction, two support vector machines (SVM) were used. The first one was trained on 50% of each cohort to distinguish TBI participants with relatively *moderate* AD-like DMN deviations from normality (i.e. with 7 or fewer statistical equivalences across TBI and AD) from other TBI participants. Another SVM was trained on 50% of each sample to distinguish TBI participants with relatively *extensive* abnormalities (i.e. with 15 or more equivalences) from the rest of the TBI participants. The means and standard deviations for the number of TNs, TPs, FNs and FPs were computed, as were their TPRs (i.e., sensitivities), TNRs (i.e., specificities), PPVs (i.e., precisions) and MCCs. Across 100 scenarios, the SVM trained to identify TBI patients whose similarities to AD were relatively modest (7 or fewer equivalences) achieved the following means and standard deviations: TN = 18.0 ± 0.8; TP = 9.0 ± 0.6; FN = 1.0 ± 0.6; FP = 1.0 ± 1.0; TPR = 0.90 ± 0.06; TNR = 0.95 ± 0.06; PPV = 0.91 ± 0.1; MCC = 0.85 ± 0.1. The SVM trained to predict which TBI patients’ similarities to AD were relatively broad (15 or more equivalences) yielded the following results: TN = 17.1 ± 0.7; TP = 10.0 ± 0.7; FN = 0.9 ± 0.7; FP = 1.1 ± 1.0; TPR = 0.9 ± 0.1; TNR = 0.95 ± 0.1; PPV = 0.9 ± 0.1; MCC = 0.86 ± 0.1. **Figure 4A** displays cortical maps of *M*_*α*_ and *M*_*β*_ on the surface of an average brain atlas, contrasting the fact that *M*_*α*_ includes primarily frontal regions whereas *M*_*β*_ includes the rest of the DMN. **Figure 4B** displays DMN parcels whose *chronic* DMN similarities across TBI and AD were accurately predicted from *acute* MoCA scores using the two SVMs. These regions include areas of dorsolateral prefrontal, lateral temporal, posterior cingulate and parahippocampal cortices.

**Figure 4.**
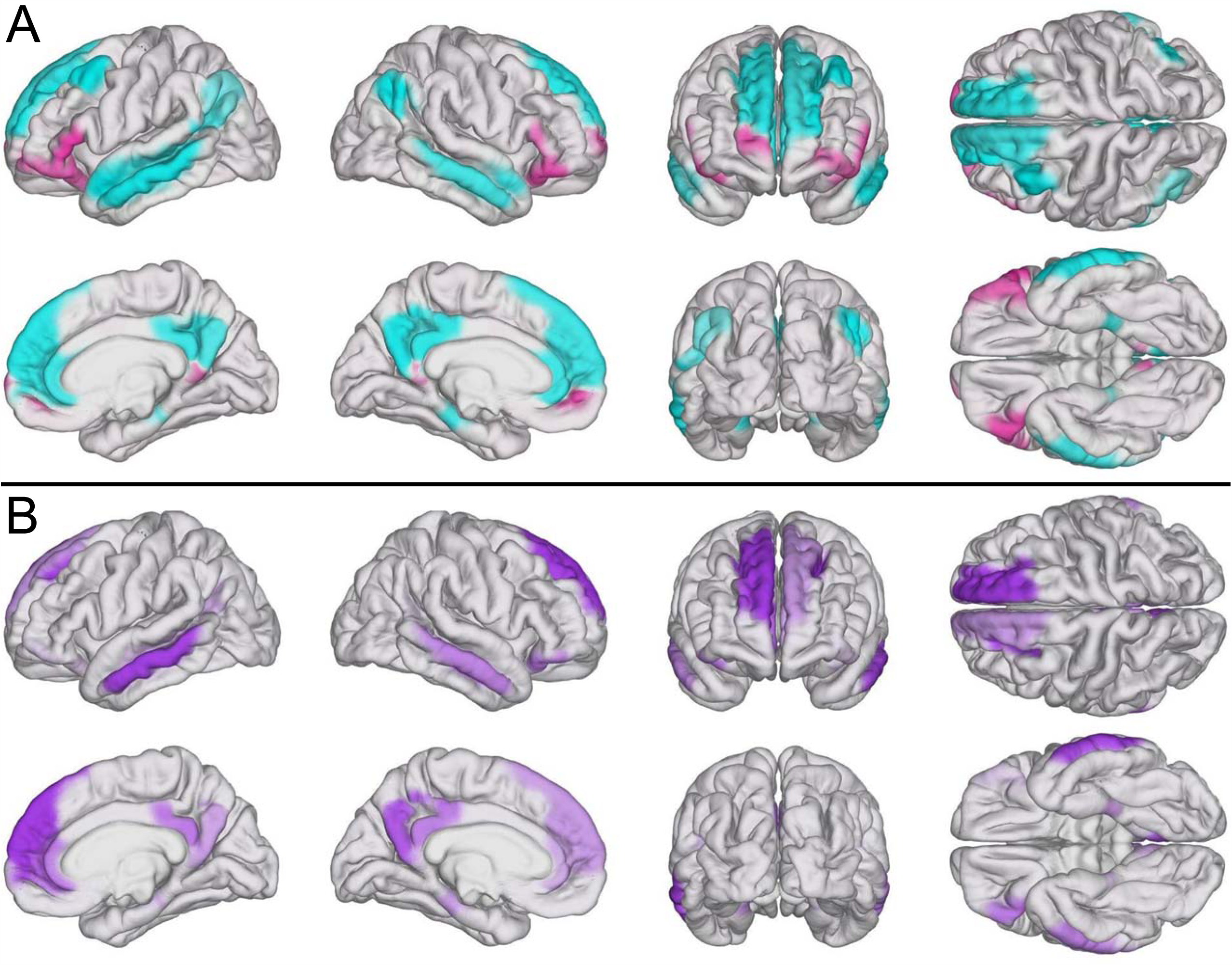
Cortical maps of DMN modules. (A) *M*_*α*_ (magenta) and *M*_*β*_ (cyan) mapped on the surface of an average brain atlas. (B) TBI-affected brain regions in *S*(*TBI, AD*) (see **Figure 3**) whose *chronic* similarity to AD was predicted based on *acute* cognitive scores using SVMs.

## Discussion

### Significance

mTBI patients with relatively high rates of neural degradation may be at commensurately high risk for AD, and the estimation of such risk can be assisted by knowledge of how TBI modifies brain function along AD-like trajectories. Thus, an important indication of this study is that the DMNs of geriatric mTBI patients can exhibit distinct patterns of AD-like RS FC as early as ∼6 months post-injury. If this is the case, our finding highlights older adults’ substantial vulnerability to TBI [32] and may be unsurprising given that the highest incidence of TBI is in older adults [5], where even injuries of mild severity can increase AD risk [8]. An alternative, more general interpretation is that geriatric mTBI patients exhibit RS FC patterns which may occur in several neurodegenerative diseases among which AD can be counted.

The outcome of the test of overall regression indicates a significant, inverse association between TBI patients’ *acute* MoCA scores and the extent of their *chronic* DMN similarities to AD. This outcome is confirmed by the SVM classifications, which achieved sensitivities and specificities which compare favorably to those for blood and imaging biomarkers of AD [33]. Our results may be useful for predicting the risk of AD-like functional degradation after TBI because mTBI patients’ acute cognitive scores are predictive of their AD-like DMN features. Thus, our study is significant in three distinct ways. Firstly, it identifies a set of functional DMN features which are common to both geriatric mTBI and AD. Secondly, it demonstrates that the analysis of RS FC in the DMN using the present approach can reveal the extent to which mTBI-affected function may transition onto AD-like trajectories. Thirdly, it suggests that, if AD-like abnormalities are indeed commensurate to mTBI patients’ AD risk, the DMN features described here can be used to improve AD risk estimation. An alternative and potentially broader implication is that our results can be used to improve risk estimation not only in AD but also in other neurodegenerative conditions whose brain degradation patterns resemble those of AD in their early stages.

### Connectomic pathophysiology

Although DMN subdivisions have been mapped with high stability across health and disease [14], such subunits’ relative vulnerability to TBI is not well understood. An intriguing finding of this study is that the node memberships of 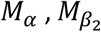 and 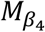 are consistently similar across the TBI and AD groups (**Figure 1** and **Figure 2**). The probability of such an occurrence being due to chance is low given the results of our randomization analysis and the fact that the modules of each dissimilarity matrix were identified independently of one another. Instead, it is more likely that this likeness of modularity could be due to intrinsic properties of the DMN which modulate their susceptibility to TBI and AD, and perhaps even to neurodegenerative processes in general.

*M*_*α*_ contains primarily fronto-hippocampo-limbic connections. For this reason, the existence of modular structure resemblances between TBI and AD may imply that both conditions impact intermodular connections within 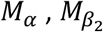 and 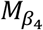 in ways which are substantially different from how they affect the rest of the DMN. Since the three modules are distinct, however, their existence may also indicate that, although FC within DMN subunits can be robust to geriatric mTBI, such robustness can manifest itself in distinct ways via mechanisms which are yet to be determined. Although appealing, the task of exploring such network-theoretic differences between DMN subunits could not be undertaken adequately here due to the small effect sizes implied by such differences. More specifically, such minute effect sizes require commensurately large samples to avoid high probabilities for errors of type I and/or II when contrasting module properties across groups. Thus, future research should aim further to study DMN similarities between TBI and AD.

The properties of the *M*_*α*_ module summarized in **Figure 1** and **Figure 2** suggest that geriatric mTBI affects certain FCs between frontal, limbic and hippocampal areas in ways which differ from those pertaining to other DMN regions. This is perhaps unsurprising given that many frontal areas experience atrophy as early as middle age [34] and that the frontal lobe appears to exhibit distinct trajectories in TBI compared to AD [35-37]. On the other hand, although FC involving the hippocampus and cingulate areas is sensitive to both TBI and AD from the early stages of both conditions [38-40], their interactions within the RS DMN remain poorly understood and require further research. Interestingly, only very few hippocampo-cortical connectivity alterations were found to be statistically similar across AD and mTBI (**Figure 3**), despite such connections being significantly affected by both conditions. This may suggest that geriatric mTBI and AD affect this connectivity ensemble in different ways, and that the functional abnormalities observed in this study might be associated with dissimilar functional trajectories within DMN modules during post-traumatic neurodegeneration. Here it is important to remind the reader of the aphorism according to which “absence of evidence is not evidence of absence.” In our context, this means that the absence of a statistical similarity finding between TBI and AD—as represented by blank connectivity matrix cells in Figure 3—does not imply evidence for the absence of such similarities. Instead, there are at least two possible scenarios. The first of these involves the situation where a similarity exists but its effect size is not large enough for its adequately powered detection to be possible in this sample. In the second scenario, although the FCs of both TBI participants and AD patients differ significantly from the FCs of HCs, the former two are not significantly equivalent. To provide an example of this, consider the scenario where the mean FC between two regions is 0 in HC, 0.8 in TBI and −0.8 in AD, and the standard deviation of each measure is 0.01. Clearly, the FC of the TBI group (0.8) and that of the AD group (−0.8) are both significantly different from that of HCs (0); nevertheless, the FCs of the TBI and AD groups differ substantially from each other (0.8 vs. −0.8), such that these FCs are not statistically equivalent. This simple example highlights the need to interpret statistical equivalence findings carefully.

In this study, the most prominent common feature of 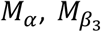 and 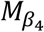 is the extent to which the intermodular connections of these three modules deviate from normality in both mTBI and AD. Furthermore, none of these modules exhibits intramodular connectivity which differs significantly from HCs in either clinical condition. These findings suggest, at the very least, that post-traumatic functional connections can be grouped according to their relative vulnerability to injury-related neurodegeneration. This is analogous to the similar task of grouping structural connections based on their vulnerability to injury, which entails mapping the structural scaffold of the human connectome [41]. Nevertheless, findings like ours may be difficult to interpret further in the absence of connectome-wide characterizations of connectivity between the DMN and the rest of the brain. Thus, future studies should aim to clarify how DMN subdivisions differ from the standpoint of their response to TBI or AD.

There are relatively few statistical similarities between TBI and AD (**Figure 3**), despite the much greater number of significant differences between HC and TBI, as well as between HC and AD (**Figure 1** and **Figure 2**). To contextualize this, it is important to note that the statistical similarities observed in this study between TBI and AD were detected only ∼6 months after geriatric TBI of mild severity. Presumably, injuries of greater severity can be associated with additional significant similarities, and future research should test this hypothesis. For example, the severity of TBI-and AD-related digestive disturbances may be associated with connectomic disruptions which affect similar cortical areas and which may be associated with injury/disease severity [42-45].

It is not unlikely that the pathophysiological processes giving rise to the relatively few observed similarities between TBI and AD continue to affect the aging brain long after injury. Thus, the number of DMN-related similarities between TBI and AD may be proportional to how late after injury such similarities are measured and quantified. Specifically, if the neuropathological processes initiated by TBI continue to affect the DMN long after injury, it is possible that TBI and AD patients’ DMNs start to resemble each another more and more as the time since injury increases. If this is the case, the number of statistical similarities observed here may provide a lower bound on the number of similarities between geriatric mTBI and AD. In such a scenario, the later fMRI recordings are acquired post-injury, the greater DMN degradation there should be along AD trajectories. Whether this conjecture is valid should be investigated by future studies.

### Neurovascular pathophysiology

The occurrence of CMBs in geriatric mTBI patients is relevant to the spatial profile and severity of post-traumatic WM alterations [16, 46, 32]. Previously, we showed that the trajectories and integrities of WM fasciculi passing through the vicinity (penumbrae) of post-traumatic CMBs can be altered in ways which persist for at least six months post-injury [47, 48]. Furthermore, other studies have found that CMB count is associated with network alterations in patients with early AD as well [49]. Since CMBs are associated with structural connectivity changes in both TBI and AD, these manifestations of blood-brain barrier breakdown may also modulate functional connectivity patterns shared by TBI and AD. In the present study, however, AD patients’ CMB counts were quite low, which suggests that CMB-related network changes previously observed in AD are relatively unlikely to drive the RS FC similarities between mTBI participants and AD patients described here. One could argue that, in ideal circumstances, (dis)similarities between HCs, mTBI patients and AD participants should be studied in the absence of CMBs; this, however, is particularly challenging in older adults. For example, in geriatric TBI patients, post-traumatic CMBs are frequently co-morbid with CMBs of hypertensive etiology, as well as with CMBS due to cerebral amyloid angiopathy (CAA), which is also a risk factor for AD. Because of the relatively high combined prevalence of neurovascular disease, CAA and hypertension in older adults [46], studying TBI-related brain network alterations in the absence of CMBs may be either logistically impractical or of limited relevance to the pathophysiological processes of the average person’s aging brain. In other words, studying functional network alterations in CMB-free older individuals may limit the utility of the insights gained from such studies to a relatively minor subset of the aging adult population. Although elucidating how CMBs modulate the extent of AD-like FC patterns in the mTBI-affected brain is beyond our scope, this study’s inclusion of individuals with a wide range of CMB counts assists in resolving AD-like FC trajectories in a sample whose neurovascular profile reflects, at least to some extent, the radiological findings of aging adults with TBI and/or AD. Future research should aim to clarify the extent to which CMBs modulate the extent and severity of AD-like FC deviations from normality in geriatric mTBI patients.

### Comparison to prior studies

Arguably, the statistical similarities between study groups (TBI and AD) are of greatest interest in this study. Nevertheless, differences between HC and TBI and between HC and AD are also relevant because they underlie key comparisons between TBI and AD [50]. Fortunately, comparison of our results to those of previous studies strengthen the case for our own analysis and broadens the scientific consensus on DMN differences between (a) HC vs. TBI and (B) HC vs. AD.

Our findings are in broad agreement with those of important previous studies on DMN abnormalities after TBI. For instance, Mayer et al. [13] reported that, compared to mTBI, HC subjects have stronger FC between (a) anterior and posterior cingulate cortices, (b) anterior cingulate cortex and the superior frontal gyrus, (c) anterior cingulate cortex and the supramarginal gyrus, (d) the inferior parietal lobule and posterior parietal cortex, (e) the inferior parietal lobule and the middle frontal gyrus, (f) prefrontal cortex and the superior parietal lobule, and between (g) prefrontal cortex and the superior frontal gyrus. Our findings are in remarkable agreement with those of Mayer et al. (**Figure 1**). Furthermore, like in the present study, Johnson et al. [11] identified stronger FC in HC participants compared to TBI volunteers between posterior cingulate cortex and the hippocampal formation. These authors also found that (a) the lateral parietal lobes have significantly more bilateral RS FCs to dorsolateral prefrontal cortex in HCs, that (b) mTBIs show only ipsilateral connections between these regions, and that (c) RS FCs between medial prefrontal and lateral parietal cortices are primarily observed in mTBI. These three sets of findings are replicated by our own study (**Figure 1**).

Influential prior results on DMN differences between HC and AD are confirmed by ours. For example, as we did, Greicius et al. [51] found that AD patients exhibit deficient activity involving the hippocampus and posterior cingulate cortex. In AD patients, Damoiseaux et al. found stronger FCs between (a) the frontal poles and other anterior frontal regions, (b) the left superior frontal gyrus and other frontal regions and between (c) the precunei and the frontal poles, but weaker FCs involving regions like the superior and middle frontal gyri. Our results confirm the findings of Greicius et al. (**Figure 2**) as well as those of Zhang et al. [52], who found reduced RS FCs between (a) posterior cingulate cortex and the hippocampus, (b) posterior cingulate cortex and the precuneus, and between (c) dorsolateral prefrontal cortex and middle temporal areas. Additionally, as we did, Zhang et al. found stronger RS FCs between the precuneus and many dorsolateral prefrontal regions.

### Equivalence testing

This study uses equivalence testing, which originates in pharmacokinetics [53]. There, marketing new drugs requires testing whether their effectiveness is undistinguishable from that of older and more expensive competitors. Here, testing whether TBI-related DMN alterations are equivalent to AD-related alterations assists in illustrating how TBI and AD can result in statistically indistinguishable patterns of DMN deviations from normality (i.e. from HCs). Using equivalence testing in studies like ours is key because attempting to establish equivalence using statistical tests of conventional null hypotheses (e.g. µ_1_ = µ_2_) frequently leads to incorrect conclusions. Specifically, a significant result after such a test establishes a difference, whereas a non-significant one simply implies that equivalence cannot be ruled out. Thus, the risk of wrongly inferring equivalence can be very high, such that proper equivalence testing is needed instead [23]. In the current context, it is important to emphasize that equivalence testing was implemented here only for pairs of regions whose partial correlations *ρ*_*ij*_ differed significantly from HCs in *both* AD *and* mTBI. If this constraint had not been imposed, distinguishing normal from abnormal statistical equivalence patterns would not have been possible within this statistical inference framework.

### Modularity structure

Here, DMN modules were identified from dissimilarity matrices rather than from FC matrices, as typical of functional connectomics studies [54]. Thus, the modules found in this study should be interpreted as groups of nodes whose FCs deviate from normality in similar ways, rather than as sets of nodes which are similarly connected to one another. The concept of deriving modularity properties from dissimilarity matrices is not new and is, in fact, the basis of multidimensional scaling (MDS)—an ordination technique for information visualization and dimensionality reduction which has been used widely for decades [55]. In MDS, like here, dissimilarity matrices can be conceptualized as distance matrices whose entries are calculated using a distance function whose definition can be conveniently assigned depending on the nature of the data. In this study, the distance in question is a *t* score, which is a proper statistical metric defined as the standardized difference between two group means. This framework is univariate and therefore accommodates only one measure at a time, i.e. functional correlation in our case. However, should additional connectivity measures (e.g. Granger causality, phase locking value) or functional modalities (e.g. electro-or magneto-encephalography) be available [56-58], this formalism could be extended to an arbitrary number of dimensions using the (multivariate) Mahalanobis distance and/or non-Euclidian metrics, like in generalized MDS [59].

### Replicability

Our findings should be replicated in larger cohorts for confirmation and improvement of statistical estimates. Although the samples used in this study were of moderate size, the effects reported here reflect relatively large mean differences between cohorts. This is perhaps unsurprising because, across a variety of studies and methodologies, even TBI of *mild* severity has been associated consistently with large statistical effects related to anatomical and physiological measures [60]. Furthermore, the mTBI participants studied here did not have clinical findings on MRI except for sporadic, SWI-detectable CMBs; this is rare in TBI studies. Thus, the present study facilitates comparison of TBI to AD partly due to the uniquely suitable profile of the geriatric TBI sample involved, whose MRI profile is relatively rare; this lends strength and uniqueness to the present study. Specifically, the effect sizes characterized here are more likely to be due to functional—rather than to structural—pathology because the structural MRI findings of these geriatric mTBI patients are minimal. For this reason, the large statistical effects of TBI upon the DMN are quite likely responsible for the large effect sizes reported. This contrasts with many other neurological conditions, where FC metrics often exhibit relatively smaller effect sizes, such that considerably larger samples are often required to detect effects of interest with adequate statistical power. It also contrasts with most other TBI studies, where gross TBI pathology on MRI findings is the norm. Nevertheless, despite the unique characteristics of our sample, further research in a larger cohort remains necessary for replication. This observation also pertains to our SVM findings, which may not be applicable to TBI cohorts of greater severity, even if only due to the greater heterogeneity of moderate-to-severe TBI relative to mTBI. Thus, our findings should not be interpreted as being broadly applicable to TBIs of any severity. Furthermore, the predictive value of our SVM relies heavily on acute MoCAs, whose values do not convey well the rich subtleties of post-traumatic cognitive impairment [61]. Thus, future studies should aim to utilize more detailed descriptors, preferably across all cognitive domains [62], to take better advantage of SVMs’ potential for functional outcome prediction. Finally, it should be mentioned that replication of our findings using electrophysiological techniques like electro-and magneto-encephalography (EEG and MEG, respectively) [63, 64] would be very helpful in establishing the spatio-temporal parameters of the (dis)similarities observed here.

### Limitations

It is important to acknowledge the possibility that the similarities between mTBI to AD described here may also be shared by mTBI with other neurodegenerative conditions. Although exploring whether this is the case is outside the scope of this study, future research should attempt to clarify whether the similarity patterns identified are representative not only of mTBI similarities to AD, but also of the relationship between mTBI and other neurodegenerative conditions like Parkinson’s disease, for which TBI is also a risk factor. Furthermore, because many participants were on medications for neurological, psychiatric, vascular, and/or metabolic disease when scans were acquired, the extent to which comorbidities affect the results of this study is unclear. Fortunately, the proportion of volunteers on medications for vascular and metabolic disease was approximately equal for the mTBI and AD groups, such that confounds due to these treatments are likely to be less severe than in the scenario where large discrepancies between groups existed. By contrast, the proportion of subjects undergoing treatment involving medications for neurological/psychiatric disease was much higher for the AD group (97%) than in the mTBI group (62%), mostly because almost all AD patients were taking cognition-enhancing medications. The effects of comorbidities upon AD-like FC trajectories in mTBI patients are worthy of further study.

One potential limitation of FC studies like ours is that results can be affected by how the DMN is defined and by the cortical parcellation used for fMRI seed analysis. Here, the DMN was defined based on the Yeo delineation and parceled based on the intersection of this delineation with the Destrieux parcellation scheme. Nevertheless, the use of other parcellation schemes of similar spatial resolution may not alter conclusions substantially because the randomization analysis undertaken yielded network modules whose anatomic coverage was consistent. Last but not least, the equivalence margin used in this study was 0.2, which is considered to be relatively conservative [23]; as the margin becomes narrower and narrower, however, more and more hypotheses of equivalence are rejected. Unfortunately, there is currently no consensus-based standard for the ‘ideal’ equivalence margin which life scientists should utilize.

## Conclusion

This study provides evidence that geriatric mTBI is associated with DMN deviations from normality which are statistically indistinguishable from those observed in AD. The DMN regions affected can be grouped into modules based on their vulnerability, with striking similarities in the composition and properties of these modules across the two neurological conditions. Multivariate regression analysis identified a clear relationship between acute cognitive deficits and chronic DMN alterations. Furthermore, SVM classifications suggested that DMN features may be useful for early prognostication of the extent and severity associated with post-traumatic neuropathophysiology. Nevertheless, the neurodegenerative processes of TBI and AD differ substantially despite their potential commonalities. Thus, the similarities in DMN alteration trajectories shared by these conditions and reported here may not be driven by similar trends toward functional reorganization. Because the methodological limitations of functional neuroimaging prevent us from a mechanistic exploration of this hypothesis, future research should study the pathophysiological mechanisms shared by TBI and AD in further detail.

## Data Availability

MRI data acquired from HC and AD participants are publicly available from the ADNI database (http://adni.loni.usc.edu). For TBI participants, primary data generated during and/or analyzed during the current study are available subject to a data transfer agreement. At the request of some participants, their written permission is additionally required in a limited number of cases.

## Acknowledgments

The authors thank Maria Calvillo, Lei Cao, Yu Hu, Jun H. Kim, Sean O. Mahoney, Van Ngo, Kenneth A. Rostowsky and Shania Wang for their assistance.

## Sources of funding

This work was supported by NIH grant R01 NS 100973 to A.I., by DoD award W81-XWH-1810413 to A.I., by a Hanson-Thorell Research Scholarship to A.I. and by the Undergraduate Research Associate Program (URAP) at the University of Southern California. Data collection and sharing for this project was funded by the Alzheimer’s Disease Neuroimaging Initiative (ADNI, NIH Grant U01 AG024904) and DoD ADNI (DoD award number W81XWH-12-2-0012). ADNI is funded by the National Institute on Aging, the National Institute of Biomedical Imaging and Bioengineering, and through generous contributions from the following: AbbVie, Alzheimer’s Association; Alzheimer’s Drug Discovery Foundation; Araclon Biotech; BioClinica, Inc.; Biogen; Bristol-Myers Squibb Company; CereSpir, Inc.; Cogstate; Eisai Inc.; Elan Pharmaceuticals, Inc.; Eli Lilly and Company; EuroImmun; F. Hoffmann-La Roche Ltd and its affiliated company Genentech, Inc.; Fujirebio; GE Healthcare; IXICO Ltd.; Janssen Alzheimer Immunotherapy Research&Development, LLC.; Johnson&Johnson Pharmaceutical Research&Development LLC.; Lumosity; Lundbeck; Merck&Co., Inc.; Meso Scale Diagnostics, LLC.; NeuroRx Research; Neurotrack Technologies; Novartis Pharmaceuticals Corporation; Pfizer Inc.; Piramal Imaging; Servier; Takeda Pharmaceutical Company; and Transition Therapeutics. The Canadian Institutes of Health Research is providing funds to support ADNI clinical sites in Canada. Private sector contributions are facilitated by the Foundation for the National Institutes of Health (www.fnih.org). The grantee organization is the Northern California Institute for Research and Education, and the study is coordinated by the Alzheimer’s Therapeutic Research Institute at the University of Southern California. ADNI data are disseminated by the Laboratory for Neuro Imaging at the University of Southern California.

## Conflicts of interest/competing interests

The authors declare no actual or perceived competing interests.

## Computer code availability

The computer code used in this study is freely available. FreeSurfer and FS-FAST are freely available (https://surfer.nmr.mgh.harvard.edu). Equivalence testing was implemented using freely available MATLAB software (https://www.mathworks.com/matlabcentral/fileexchange/63204). Network analysis was implemented using the freely available Brain Connectivity Toolbox (https://sites.google.com/site/bctnet/). Network visualizations were generated using Gephi (http://gephi.org). Regression and SVM analyses were implemented in MATLAB (http://mathworks.com) using the `glmfit, fitcsvm` and `predict` functions.

## Authors’ contributions

A.I. contributed to study design, data analysis, result interpretation and wrote the manuscript. A.S.M., N.N.C., N.F.C. and E.B.J. and contributed to study design, data analysis and result interpretation.

## Notes

### Competing Interest Statement

The authors have declared no competing interest.

### Author Declarations

This study was conducted with the approval of the Institutional Review Board at the University of Southern California.

